## Supplementary material for "Systematic evaluation of multifactorial causal associations for Alzheimer’s disease and an interactive platform based on Mendelian randomization analysis——MRAD": Certificate_of_editing-OMQJC_10_1tcin2wx-d.pdf

### CERTIFICATE OF ENGLISH EDITING

This document certifies that the paper listed below has been edited to ensure that the language is clear and free of errors. The edit was performed by professional editors at Editage, a division of Cactus Communications, in cooperation with Taylor & Francis Group. The intent of the author's message was not altered in any way during the editing process. The quality of the edit has been guaranteed, with the assumption that our suggested changes have been accepted and have not been further altered without the knowledge of our editors.

#### Title

Systematic evaluation of multifactorial causal associations for Alzheimer's disease and an interactive platform based on Mendelian randomization analysis——MRAD

#### Authors

Tianyu Zhao1 , Hui Li2,3 , Meishuang Zhang4 , Yang Xu1 , Ming Zhang1 , Li Chen1*

#### Order No.

OMQJC_10

EDITINGSERVICES
Supporting Taylor & Francis authors

Signature

Vikas Narang

Vikas Narang,
Chief Operating Officer,
Editage

Date of Issue
**April 07, 2023**

editage

**Taylor & Francis Editing Services**

www.tandfeditingservices.com
