## Supplementary material for "Systematic evaluation of multifactorial causal associations for Alzheimer’s disease and an interactive platform based on Mendelian randomization analysis——MRAD": SUPPLEMENTARY MATERIAL.docx

**SUPPLEMENTARY MATERIALS**

**MRAD User Guide**

**Home module** (as shown in Figure S8)

The Home module allows users to search through 400,274 records. Users can enter search terms, such as exposure (e.g., LDL cholesterol) or id.exposure (e.g., ebi-a-GCST000759), in the search box. The search interface includes 20 control widgets, as shown in Figure S9. Among these control widgets, there are eight select boxs, including Classification I, Classification II, Classification III, id.outcome, method, effect_direction, Consortium, and Sex, allowing users to make single or multiple selections to filter the data. The control widgets also include two text input boxs, id.exposure and exposure, which allow users to search by text input (case-insensitive). The control widgets Min pval and Max pval allow users to search by numerical input (initial values: Min pval = 0, Max pval = 1). The Heterogeneity test and MR-Egger intercept test checkboxes provide users with a single-click option to obtain data results that have passed heterogeneity and horizontal pleiotropy tests (p>0.05). The Year range slider helps users to filter data based on the year of publication (range: 2007-2022). Additionally, we have supplemented relevant basic information on biomarker functions and pathways from the public database Uniprot. Users can search for information using the Uniprot Entry ID and Gene Names input boxs and can click on the Uniprot Entry ID Link in the table to go to the corresponding Uniprot page. The Download, Reset, and Back buttons respectively provide users with the ability to download the data in a .csv format, reset filter conditions of all control widgets, and return to the Home interface.

Furthermore, in the Home module, users can also access the 10 category results of the IVW model (the major analysis method), by checking the corresponding checkbox on the right-hand side (note: only one checkbox can be checked at a time). For example, checking the "Disease" checkbox will allow users to jump to the results page containing 17,168 disease records.

**Study Design module**

The Study Design module provides users with a complete MR study design. Moreover, corresponding checkboxes are provided below to allow users to easily access the corresponding details (note: only one checkbox can be selected at a time), as illustrated in Figure S10.

**IVW interactive and IVW static modules**

The IVW interactive module, as shown in Figure S11, contains 21 control widgets, including the same search control widgets as in the Home module (due to the large amount of data, the initial values for Classification I and Classification II are set to "Medical laboratory science" and "blood lipids and lipoproteins", respectively; users can reset the conditions as needed). In addition, the IVW interactive module includes the following new features: (i) a checkbox for "Exposures with no effect", which, when selected (checked by default), allows users to simultaneously remove all exposure traits that do not have significant association (p>0.05) with any of the 16 outcome traits; (ii) a checkbox for "80 traits with consistent effect across three main outcome datasets", which, when selected (unchecked by default), provides users with the option to filter results that have significant and consistent causal association with all the three main outcome traits of AD (p<0.05); (iii) Download Data and Download Interactive buttons provide users with the ability to download data in a .csv format and images in a .html format, respectively.

The IVW interactive module also provides users with interactive visual results (Figure S12). Clicking on the dots on the image will display detailed information on the corresponding id.exposure, exposure, id.outcome, outcome, beta, OR_CI, pval, ‑log10(pval), nsnp, method, Heterogeneity test, and MR-Egger intercept test.

The IVW static module is the same as the IVW interactive module in terms of all the control widgets except for the absence of the Download Interactive button, and contains only static graph results.

**Sensitivity analysis interactive and Sensitivity analysis static modules**

The Sensitivity analysis interactive module contains 21 control widgets, all of which are the same as those in the IVW interactive module, except that the "80 traits with consistent effects across three main outcome datasets" checkbox is absent, and that the id.outcome option is only available for single selection (initial value: ieu-b-2, users can reset the conditions as needed). In addition, a Sensitivity Analysis Passed select box widget has been added, which allows users to select one or more of the six sensitivity analysis models and obtain statistically significant results (p<0.05), as shown in Figure S13. Interactive visual results are also provided to users. By clicking on the squares in the image, detailed information for id.exposure, exposure, id.outcome, outcome, beta, OR_CI, pval, nsnp, and method is displayed, as shown in Figure S14. The Sensitivity Analysis Static module has the same control widgets as the Sensitivity Analysis Interactive module, except that it does not include the Download Interactive button and only contains static image results.

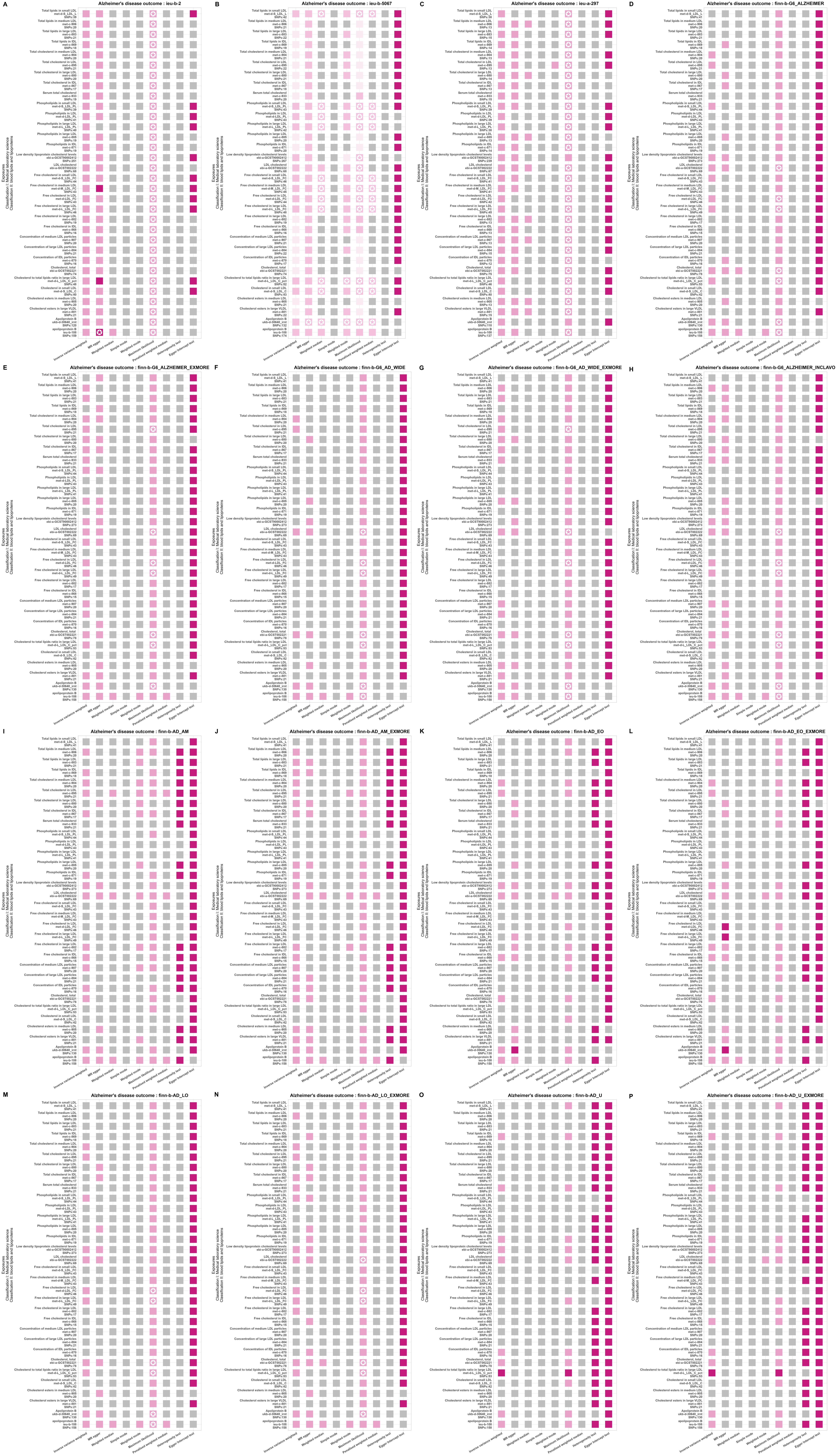

Figure S1. Statistical models for causal effect results of thirty-two blood lipids and lipoproteins items that were positively associated with the main outcome traits of AD.

Note:

(i) For column Inverse variance weighted, MR egger, Weighted median, Simple mode, Weighted mode, Maximum likelihood, and Penalized weighted median: the pink dots in the figure represent positive association, the blue dots represent negative association, with the color depth of the dots being positively proportional to the OR value (the darker the color, the larger the OR value), and the size of the dots being inversely proportional to the p-value (the smaller the p-value, the larger the dots). The gray dots represent no significant causal association (p>0.05). The star mark(✪) represents that is significant at the Bonferroni threshold (p＜1.727e-07).

(ii) For column Heterogeneity test: the pink dots in the figure represent the effect of heterogeneity was considered negligible (heterogeneity_pval> 0.05). The gray dots represent significant association (p＜0.05).

(iii) For column Egger intercept test: the pink dots in the figure represent there was no significant difference between Egger Intercept and 0, indicating no horizontal pleiotropy (Horizontal_pval> 0.05). The gray dots represent significant association (p＜0.05). The dark gray dots represent not applicable due to the quantity of SNP was less than 3.

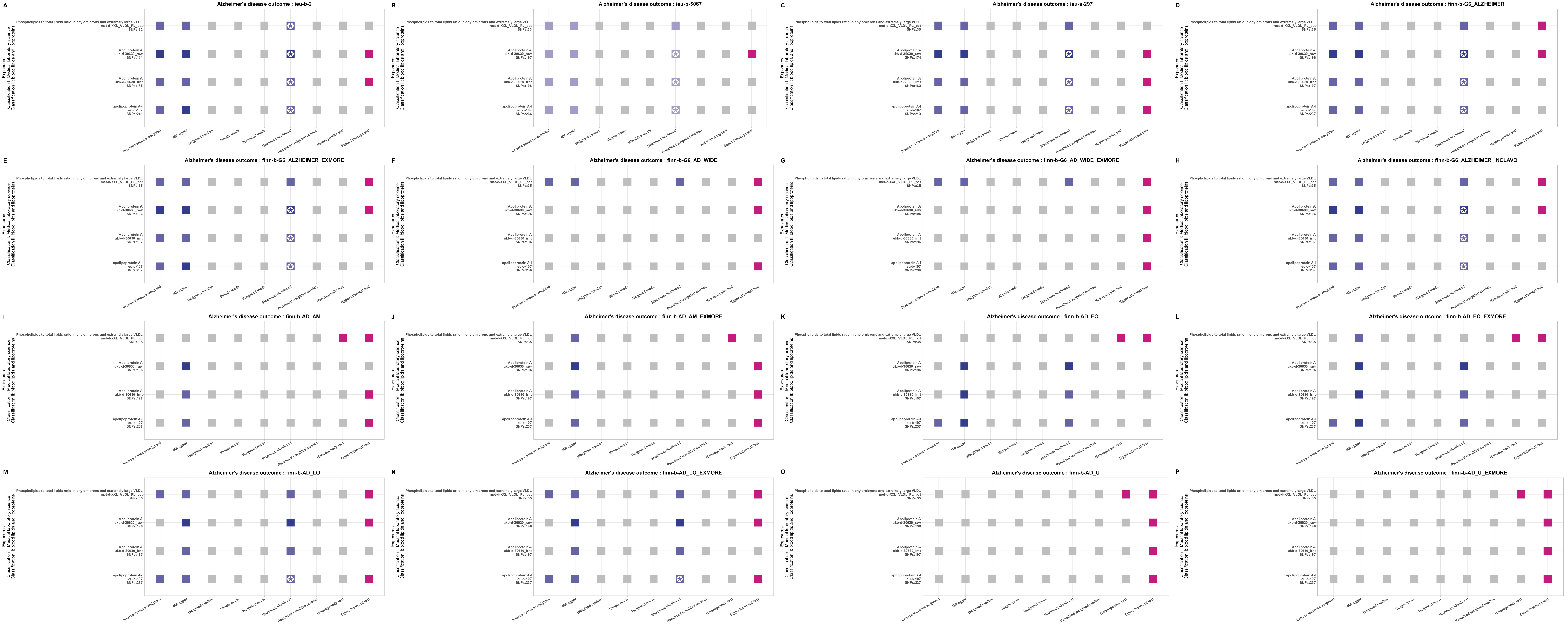

Figure S2. Statistical models for causal effect results of four blood lipids and lipoproteins items that were negatively associated with the main outcome traits of AD. The figure note follows that of Figure S1.

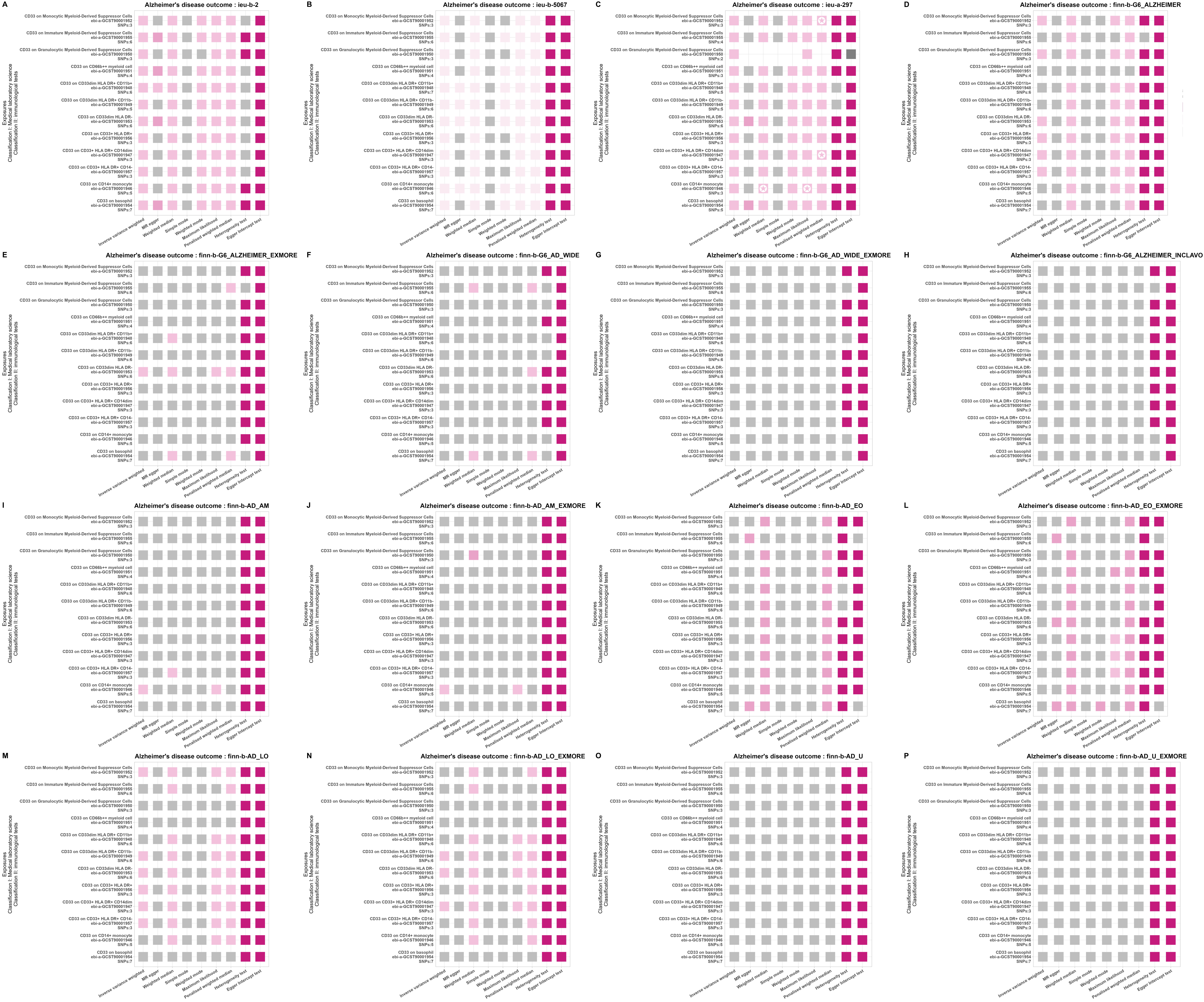

Figure S3. Statistical models for causal effect results of twelve immunological test items that were positively associated with the main outcome traits of AD. The figure note follows that of Figure S1.

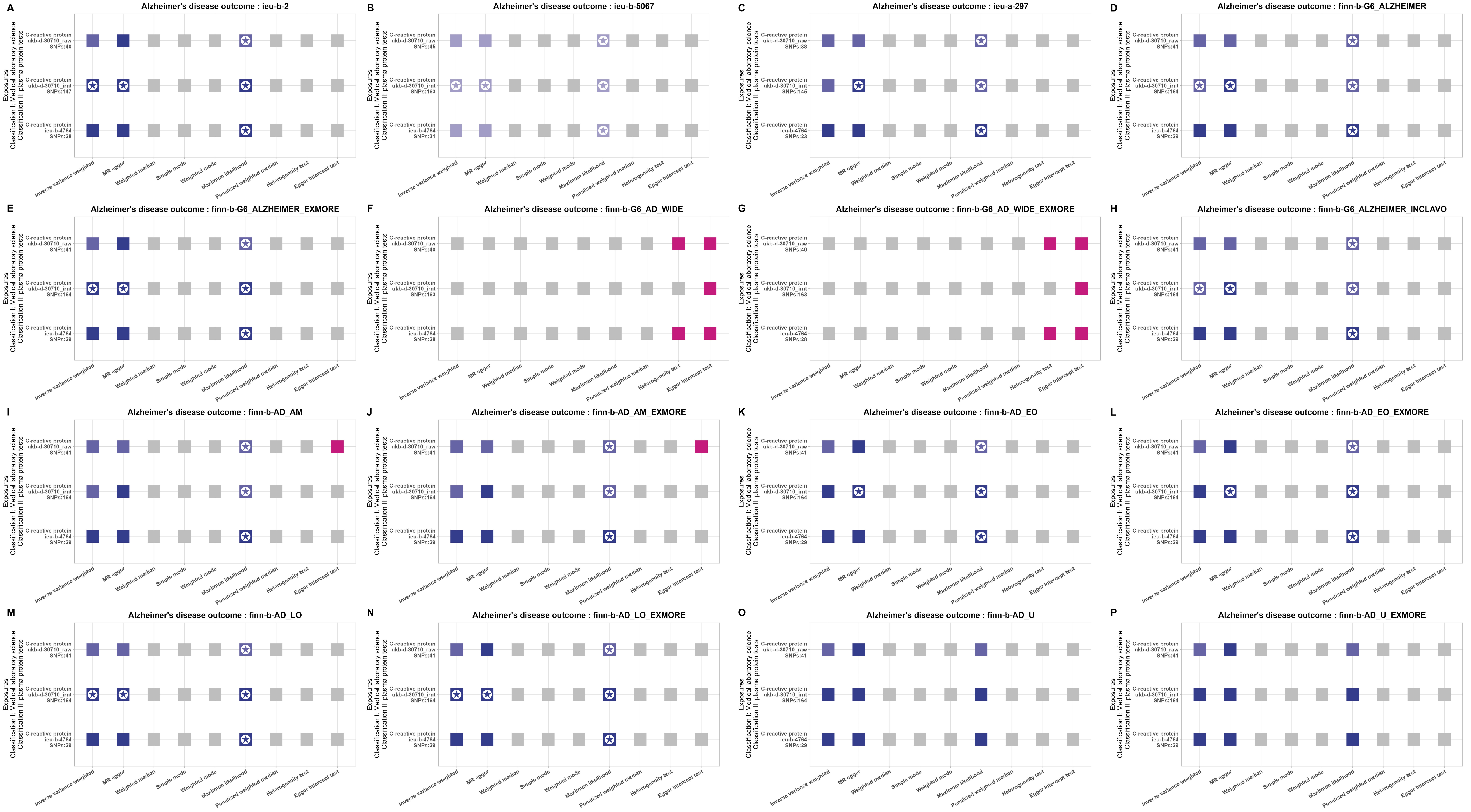

Figure S4. Statistical models for causal effect results of three plasma protein tests items that were negatively associated with the main outcome traits of AD. The figure note follows that of Figure S1.

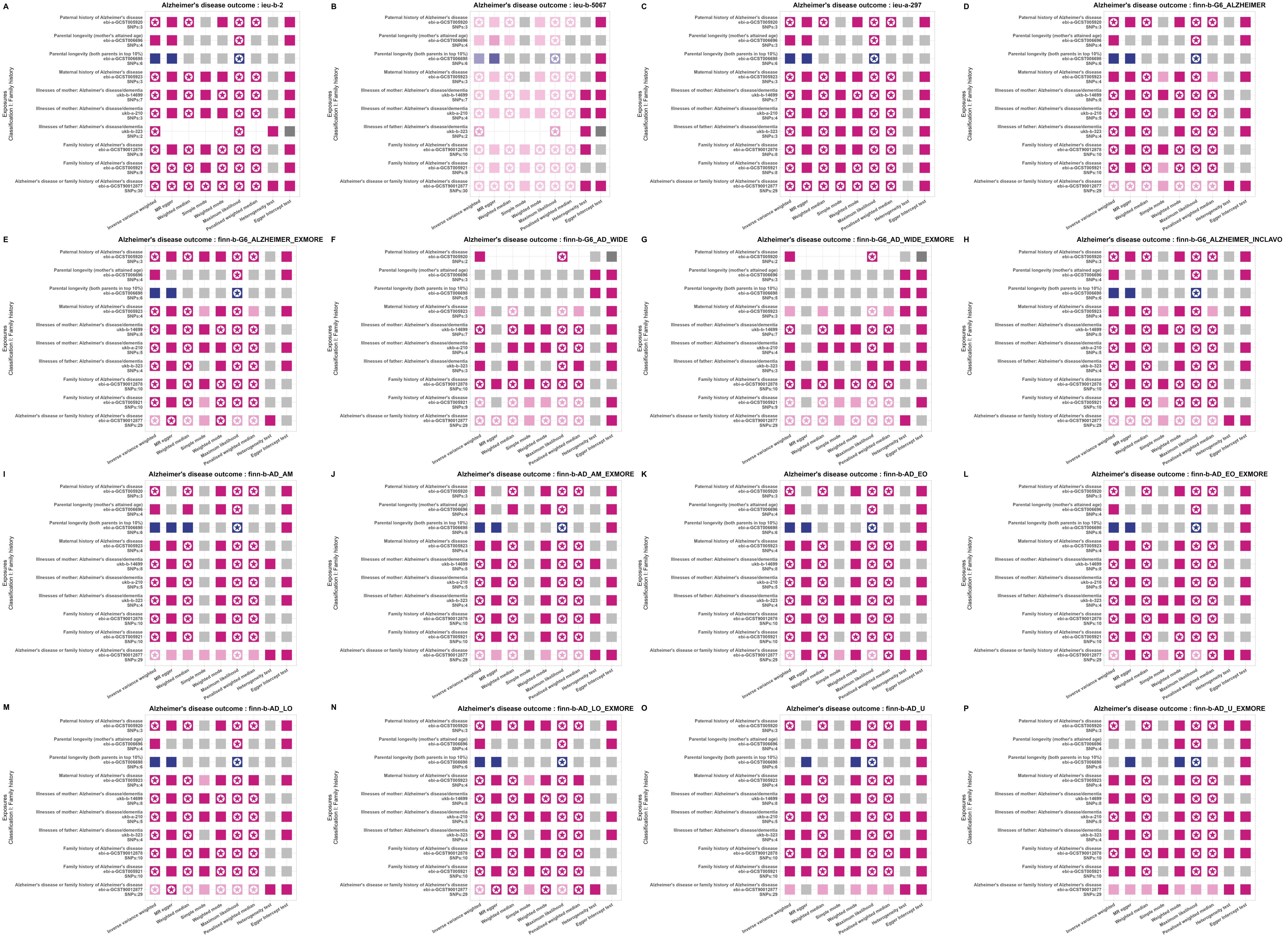

Figure S5. Statistical models for causal effect results of ten family history items with causal effects on the main outcome traits of AD. The figure note follows that of Figure S1.

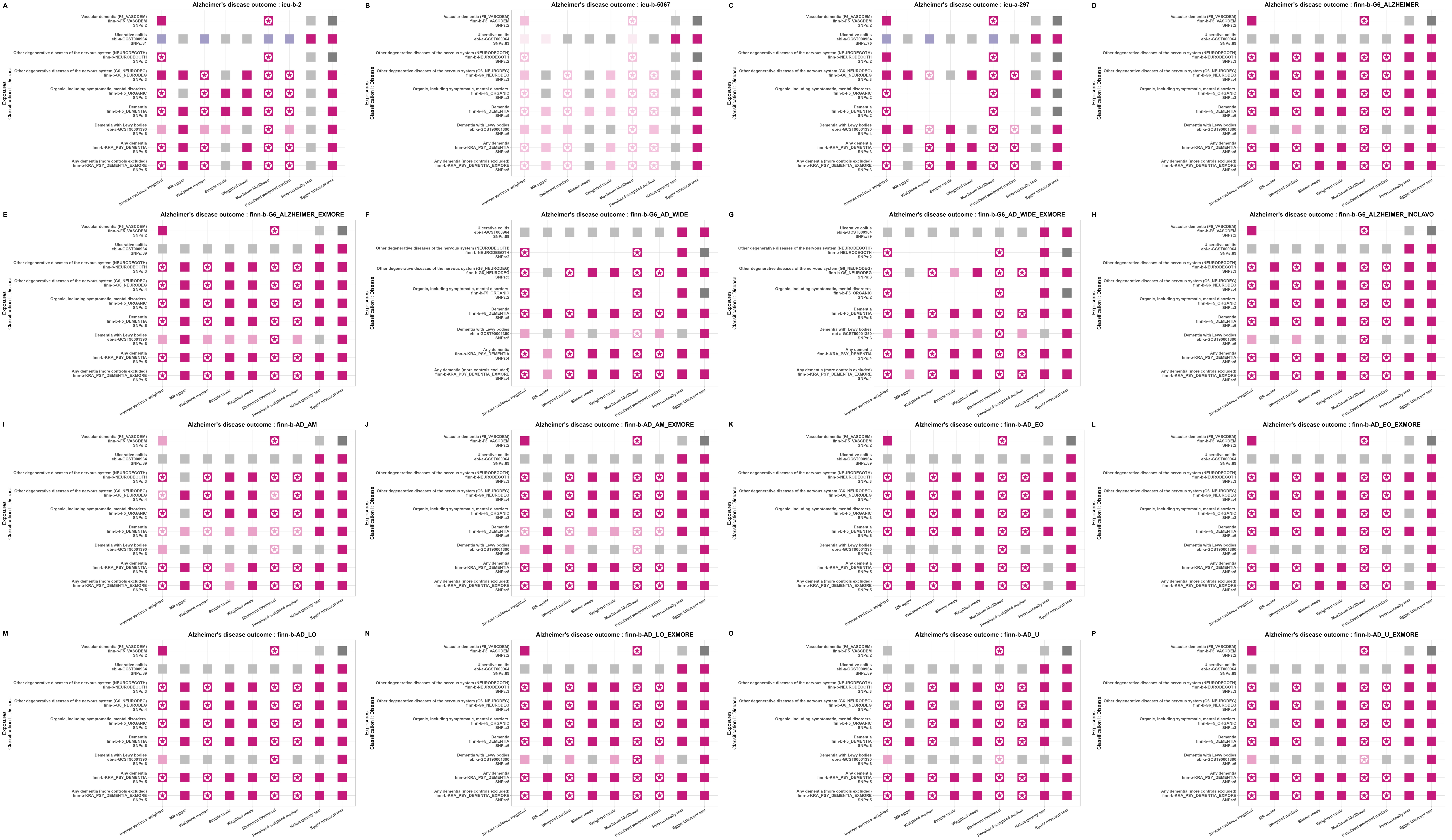

Figure S6. Statistical models for causal effect results of nine diseases items with causal effects on the main outcome traits of AD. The figure note follows that of Figure S1.

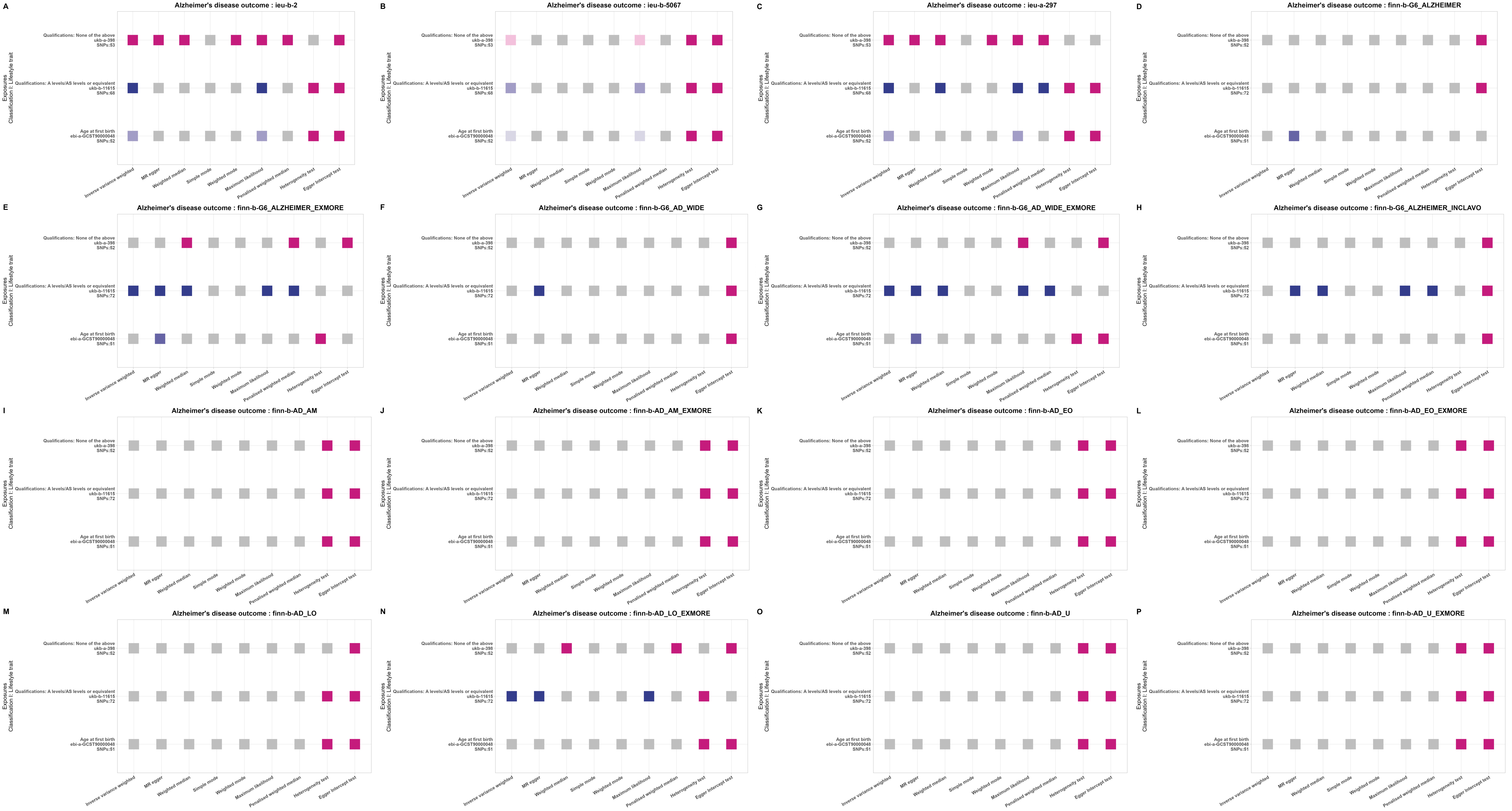

Figure S7. Statistical models for causal effect results of three lifestyle trait items with causal effects on the main outcome traits of AD. The figure note follows that of Figure S1.

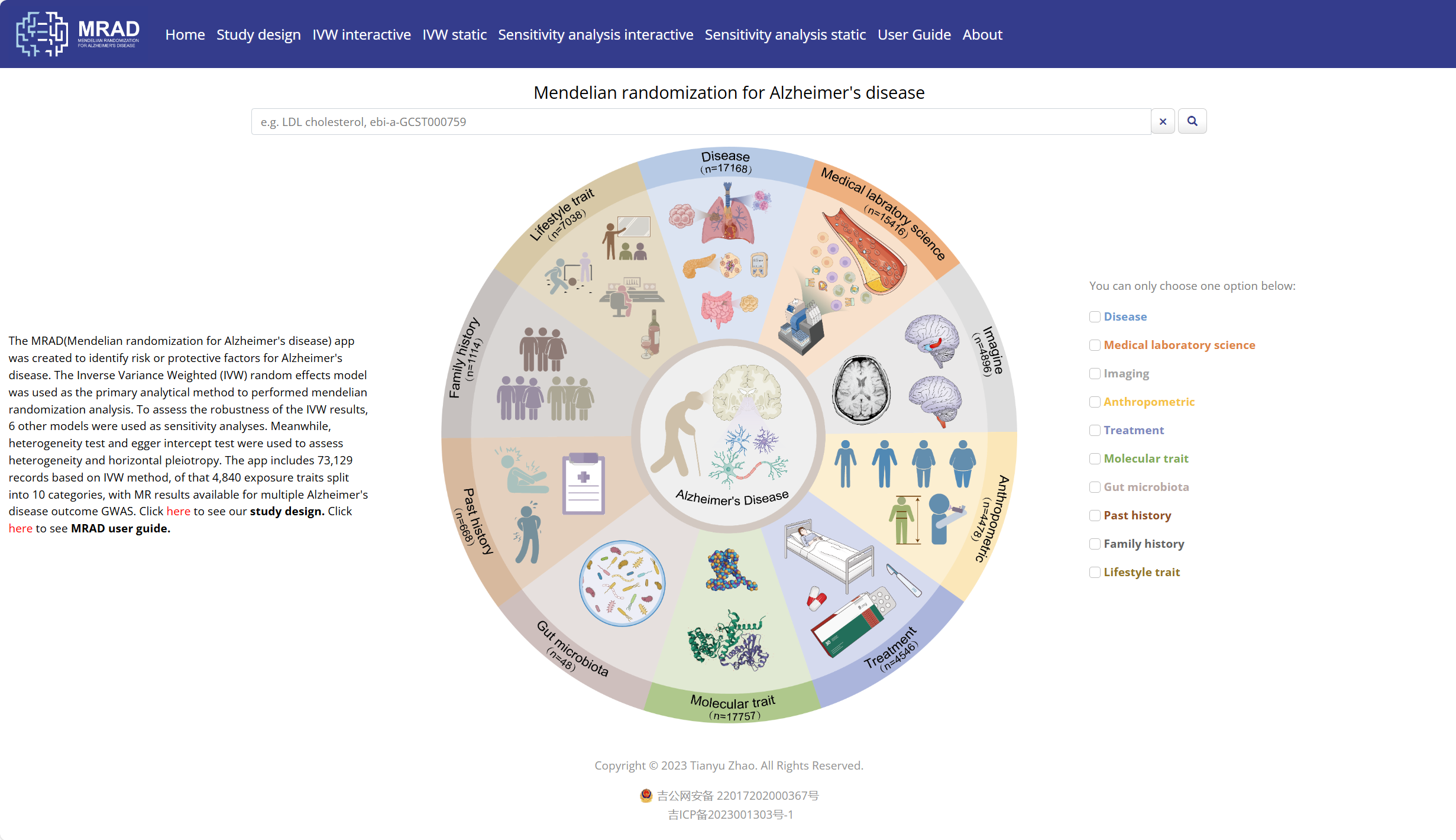

Figure S8. Home module

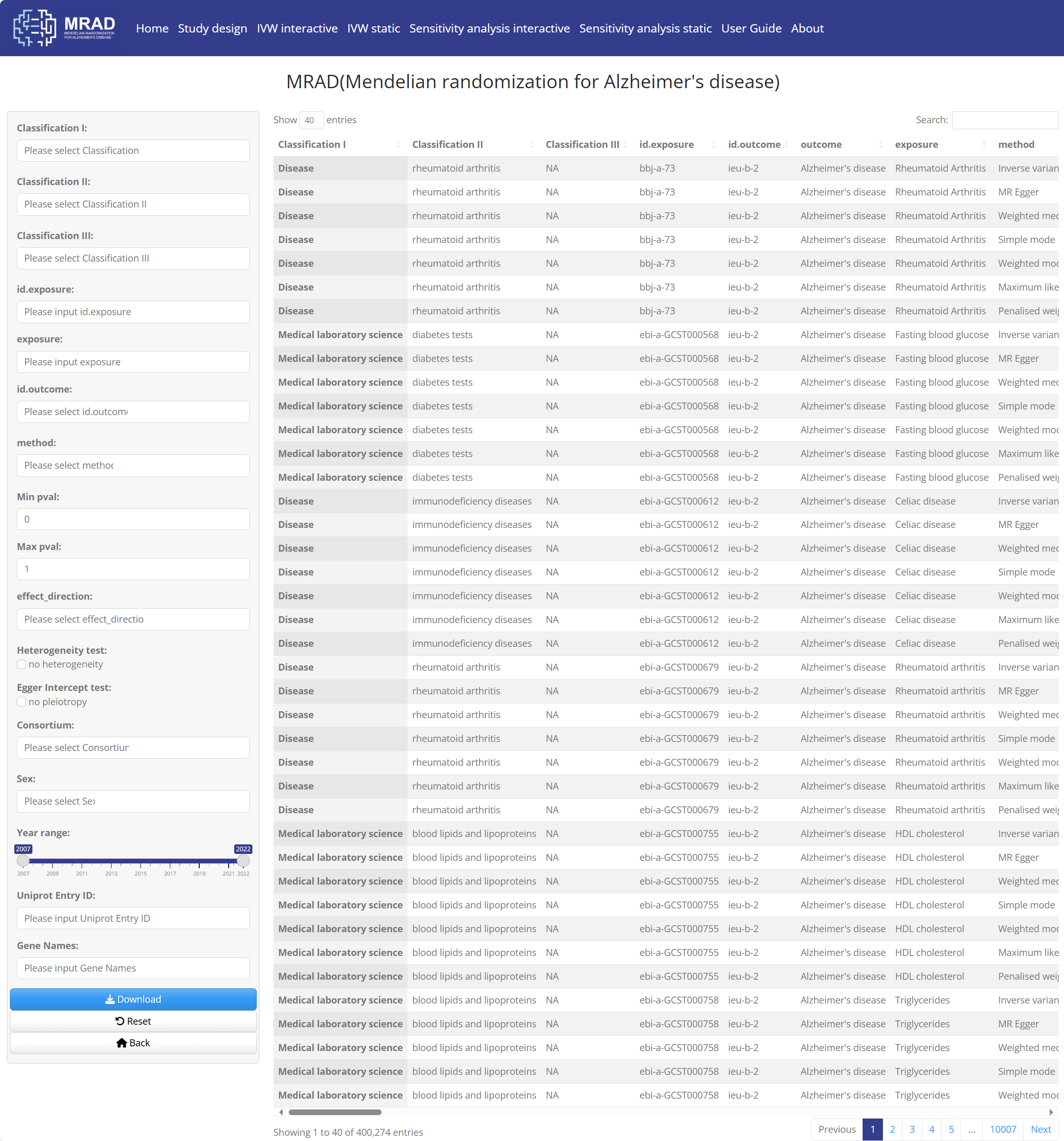

Figure S9. 400,274 records search interface of MRAD

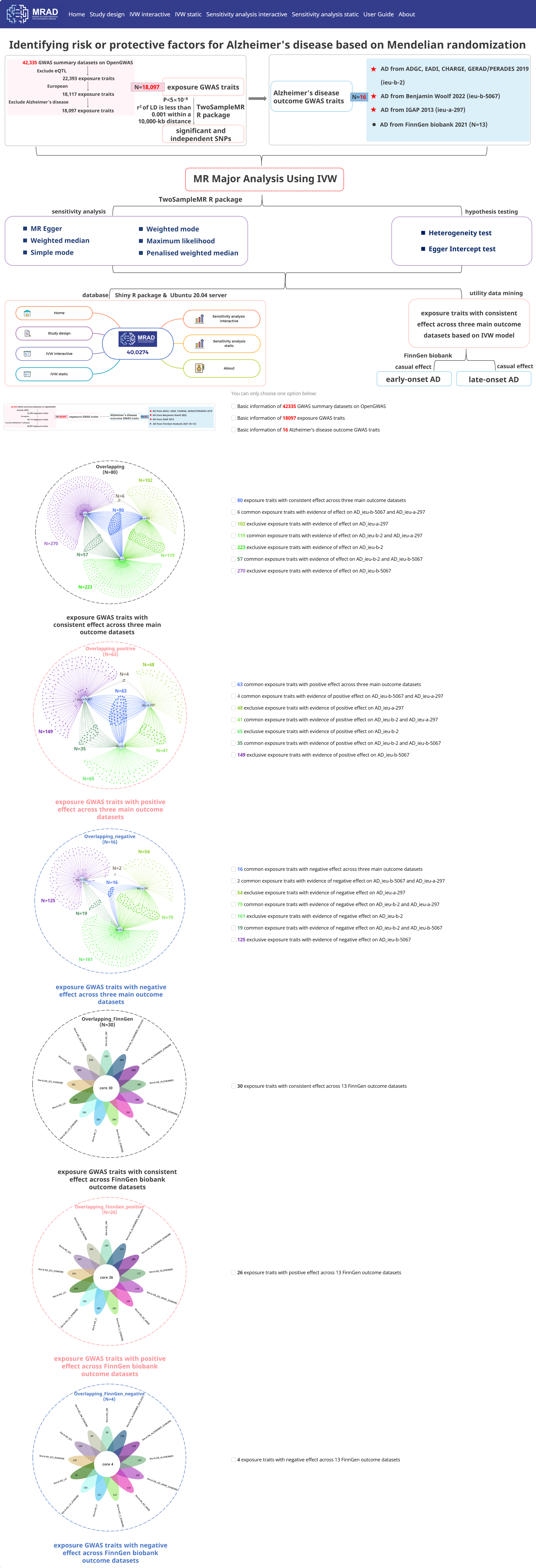

Figure S10. Study Design module

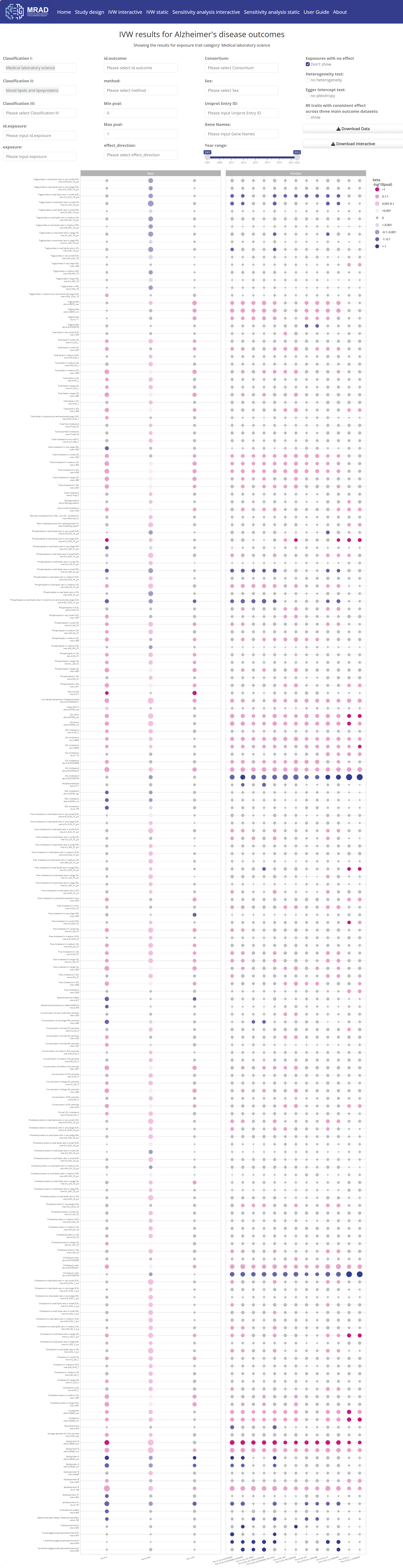

Figure S11. IVW interactive module

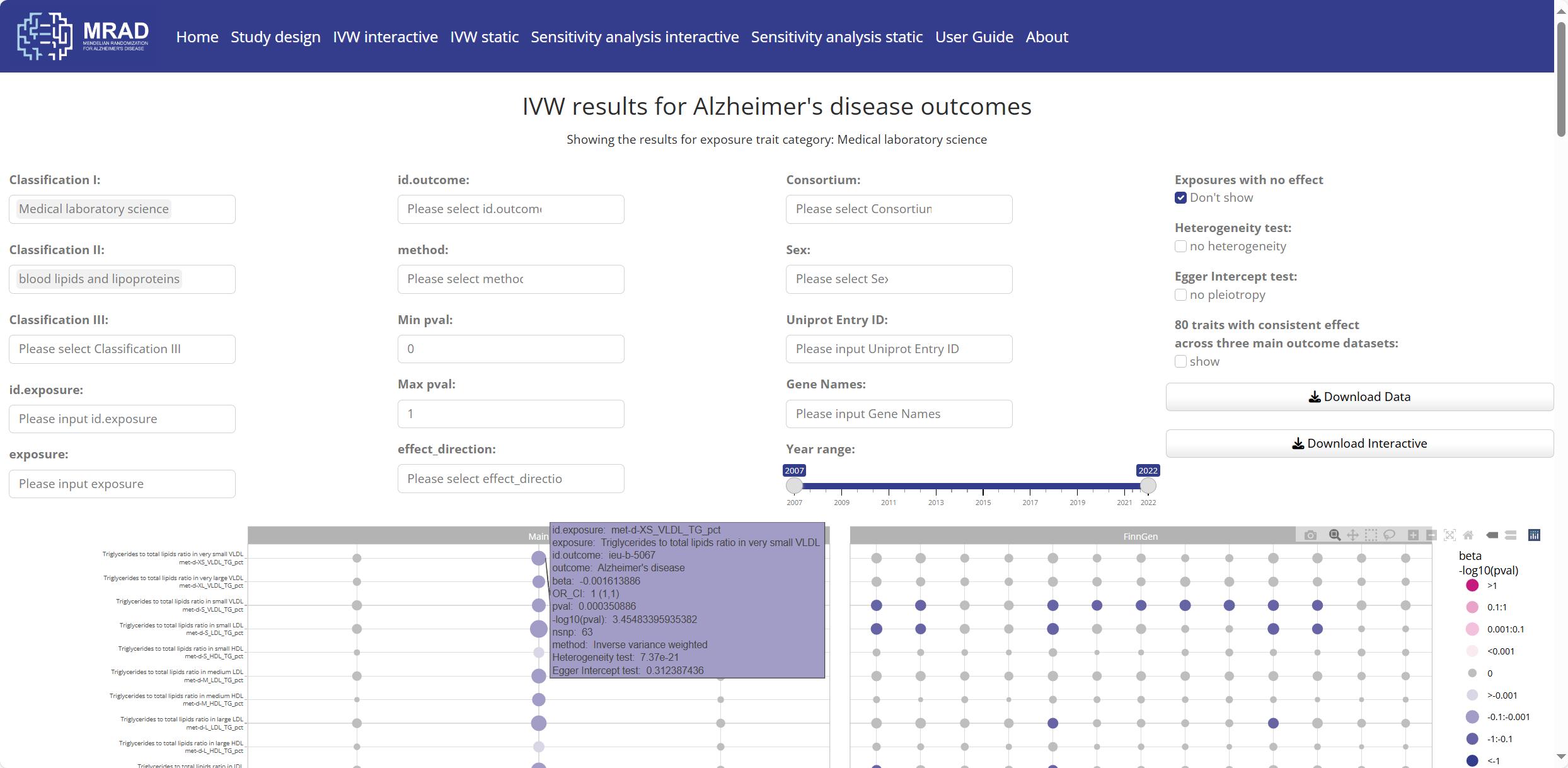

Figure S12. Interactive visual results in the IVW interactive module

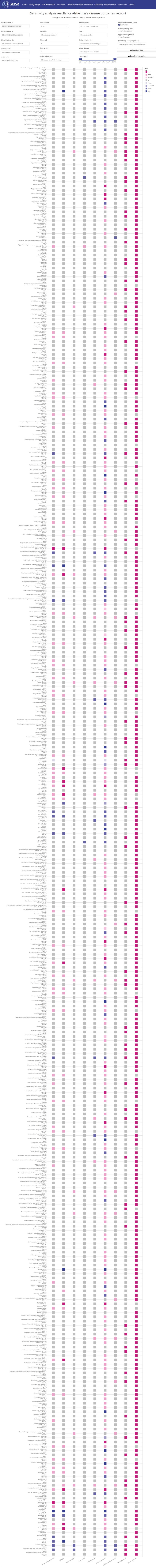

Figure S13. Sensitivity analysis interactive module

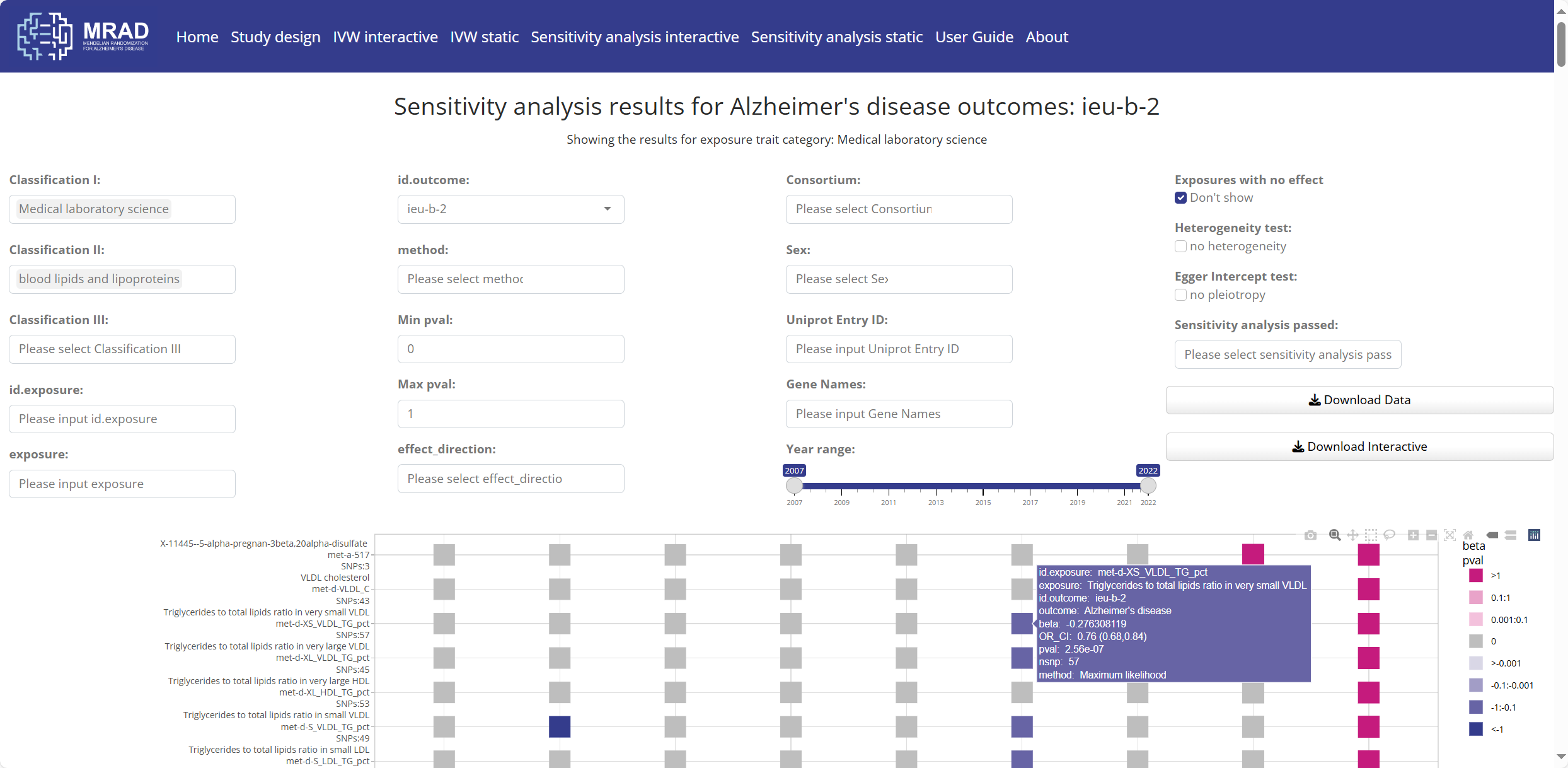

Figure S14. Interactive visual results in the Sensitivity analysis interactive module
